# RBDAct: Home screening of REM sleep behaviour disorder based on wrist actigraphy in Parkinson’s patients

**DOI:** 10.1101/2022.01.23.22269713

**Authors:** Flavio Raschellà, Stefano Scafa, Alessandro Puiatti, Eduardo Martin Moraud, Pietro-Luca Ratti

**Author notes:** These authors contributed equally to this work (Eduardo Martin Moraud, Pietro-Luca Ratti). **Correspondence to** Pietro Luca Ratti, Neurocenter of Southern Switzerland, Ente Ospedaliero Cantonale, via Tesserete, 46, CH-6903 Lugano, Switzerland.

## Abstract

**Background:** REM sleep behaviour disorder (RBD) is a disabling, often overlooked sleep disorder affecting up to 70% of patients with Parkinson’s disease. Identifying and treating RBD is critical to prevent severe sleep-related injuries, both to patients and bedpartners. Current diagnosis relies on nocturnal video-polysomnography, which is an expensive and cumbersome exam requiring specific clinical expertise.

**Objectives:** To design, optimise, and validate a novel home-screening tool, termed RBDAct, that automatically identifies RBD in Parkinson’s patients based on wrist actigraphy only.

**Methods:** Twenty-six Parkinson’s patients underwent two-week home wrist actigraphy worn on their more affected arm, followed by two non-consecutive in-lab evaluations. Patients were classified as RBD versus non-RBD based on dream enactment history and video-polysomnography. We characterised patients’ movement patterns during sleep using raw tri-axial accelerometer signals from wrist actigraphy. Machine learning classification algorithms were then trained to discriminate between patients with or without RBD using actigraphic features that described patients’ movements. Classification performance was quantified with respect to clinical diagnosis, separately for in-lab and at-home recordings.

**Results:** Classification performance from in-lab actigraphic data reached an accuracy of 92.9±8.16% (sensitivity 94.9±7.4%, specificity 92.7±13.8%). When tested on home recordings, accuracy rose to 100% over the two-week window. Features showed robustness across tests and conditions.

**Conclusions:** RBDAct provides reliable predictions of RBD in Parkinson’s patients based on home wrist actigraphy only. These results open new perspectives for faster, cheaper and more regular screening of sleep disorders, both for routine clinical practice and for clinical trials.

## INTRODUCTION

REM sleep behaviour disorder (RBD) is a sleep disorder affecting up to 70% of patients with Parkinson’s disease (PD)^1^. Patients with RBD exhibit movements and dream enactment behaviours during sleep which can be vigorous, sometimes violent and harmful^2^. Diagnosing and treating RBD is of pivotal importance to prevent severe injuries to patients and their bedpartners.

Isolated RBD represents an early stage of PD or other synucleinopathies^3^, and can precede for several years more overt clinical manifestations of these disorders^4 5^. Its early diagnosis offers a unique window to evaluate disease-modifying effects of upcoming treatments^6^. Additionally, PD phenotypes that are associated with RBD tend to be more aggressive and to exhibit more motor complications. They are also more often accompanied by cognitive, behavioural and dysautonomic symptoms^7^. Identifying RBD in PD can thus provide fundamental insights to inform clinical practice, both from a therapeutical and prognostic point of view^8^.

RBD remains an overlooked and underrecognized phenomenon even among movement disorders specialists. RBD diagnosis requires nocturnal video-polysomnography (VPSG)^2^, which is a costly, time-consuming exam that is only accessible in specialised centres and can be burdensome for patients.

Current screening tools rely on questionnaires or interviews. However, these approaches are often subjective, and can either not be available for community-dwelling individuals^9^ or require the presence of a bedpartner^10^. In Parkinson’s patients, their reliability to capture RBD is not well established^11-13^. Little progress has been made in the development of objective screening tools for RBD diagnosis in everyday life settings. This would be a mainstay to better understand RBD manifestations and their changes over time, and to assess treatment efficacy during clinical trials and clinical routine^6^.

In this study, we designed and validated a novel, wearable approach for identifying RBD automatically at home in PD patients. We combined actigraphic technology and state-of-the-art machine learning algorithms that were optimised in controlled clinical settings and translated to home environments.

## PATIENTS AND METHODS

### Study design and population

#### Ethical considerations

The study was conducted in the framework of the *Awake & Move study*^14 15^. It was approved by the Ethics committee of the Canton of Ticino, Switzerland (Ref. 2016-00056) and conducted in accordance with the Declaration of Helsinki. Written consent was provided by all participants. Participation in this study was on a voluntary basis and proposed to all patients meeting the eligibility criteria who were attending the outpatient department of the Movement Disorder Unit of the Neurocenter of Southern Switzerland in Lugano, Switzerland. Additional patients volunteered to participate after advertisements in the magazine of the Swiss Parkinson’s association, and in public conferences organised by the same association.

#### Inclusion and exclusion criteria

Eligibility criteria were: mild to moderate idiopathic PD (no atypical parkinsonism)^16^ (Hoehn & Yahr stage >1 and ≤3)^17^, no cognitive impairment (Mini-Mental State Examination score ≥26/30)^18^, no active depression (Beck Depression Inventory score < 14/63)^19^, no deep brain stimulation.

### Study procedures

#### Patients’ participation and workload

An initial recruitment visit (V0) was organised at the hospital by a senior neurologist, expert in sleep medicine and movement disorders, who performed a thorough medical and neurological examination. Evaluations included sleep history and the Movement Disorders Society Unified Parkinson’s Disease Rating Scale (MDS-UPDRS), with the motor part (III) performed during the “on” phase in patients with motor fluctuations.

In each recruited patient, sleep and wake patterns were profiled by means of continuous actigraphy monitoring, recorded at home over a 2-week period, coupled with an electronic sleep diary. Sleep and wake routines were recorded by means of a proprietary application for tablets, *SleepFit*^20^.

At the end of this period, a full in-lab video-polysomnography (VPSG) was performed. The times of “lights-out” and “lights-on” were set for each subject according to their usual bed-and wake-time schedules, mirroring sleep habits of the previous 2 weeks. Habitual hypnotic medications and other psychotropic agents were allowed during the subjects’ participation in the study. Alcoholic, caffeinated or other stimulant beverages, as well as tobacco smoking, were not permitted 4 hours prior to bedtime. A second VPSG was performed 7 to 14 days after the first one. Between the first and the second VPSG recordings, the patients were asked to keep their routines and daily medications unchanged.

#### Wrist actigraphy

*GENEActiv Original* wrist actigraph (GENEActiv™, Activinsight Ltd., Kimbolton, Cambridgeshire, UK)^21^, worn on the more affected arm, was employed during the 2-week home recordings. It recorded tri-axis arm accelerations (*a*_*x*_, *a*_*y*_, *a*_*z*_) and environmental light. Signals were acquired at 40 Hz sampling frequency, to maximise battery duration. In parallel to the in-lab VPSG recordings, continuous recordings of motor activity were acquired using the same *GENEActiv Original* devices, set to record at a 100-Hz sampling frequency, and worn on both wrists.

#### Video-polysomnography

VPSG recordings were performed according to the American Academy of Sleep Medicine standards^22 23^, including scalp electroencephalography, electro-oculogram, surface electro-myogram of the chin, lower limbs electromyograms^24-26^, nasal and oral flow, respiratory effort sensors, pulse oximeter and electrocardiogram. Synchronised digital infrared video tracks and ambient sound recordings were also acquired (**Fig. 1**). Visual analysis of PSG recordings were performed by a trained sleep and movement disorder expert (PLR) according to standard criteria^22 23^, taking into account previously published recommendations and suggestions for sleep scoring in PD^2 27^.

**Figure 1.**
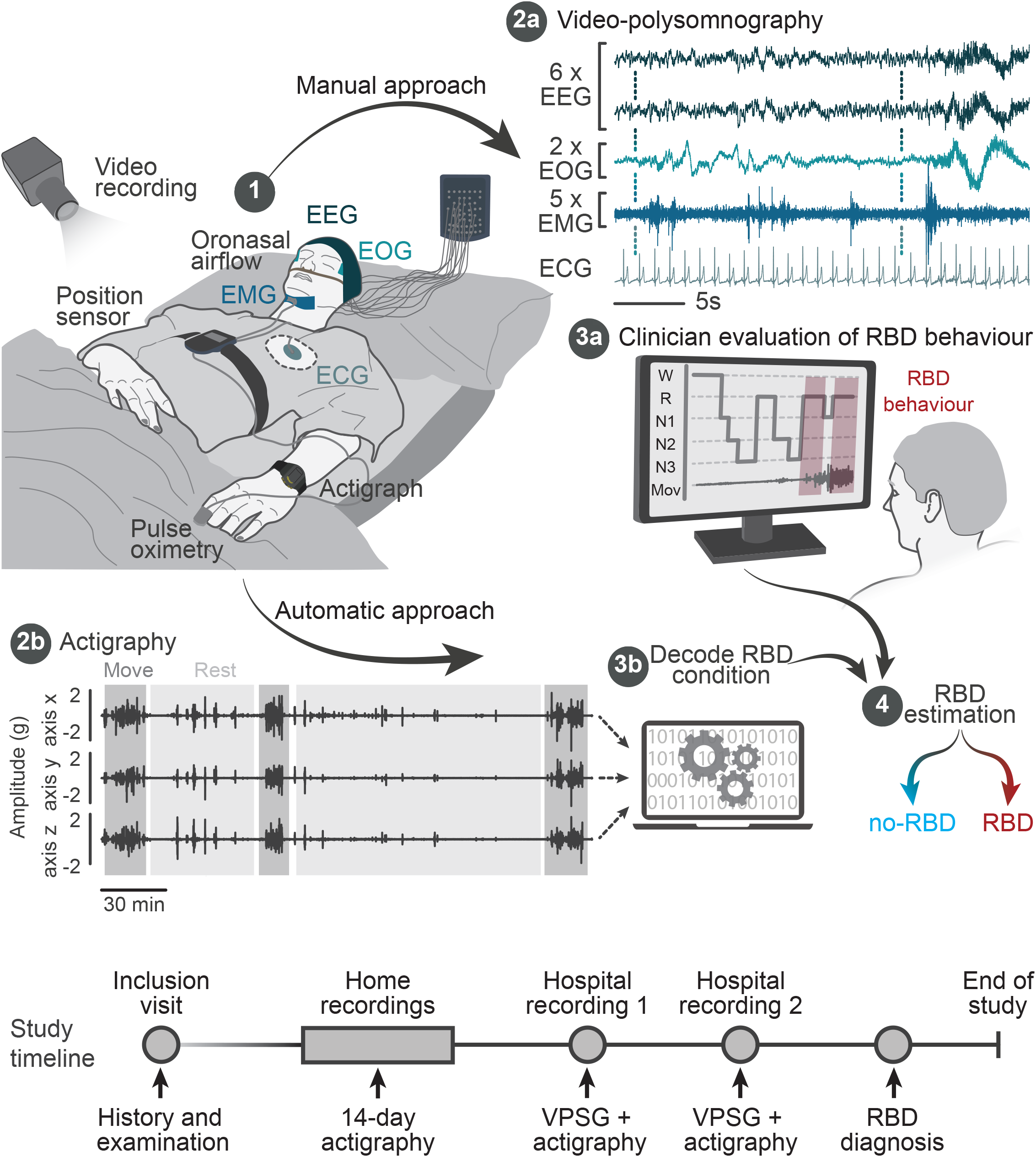
Experimental setup for in-lab recordings and study design. Video-polysomnography (VPSG) was recorded concurrently to actigraphy (1). All VPSG signals were displayed (2a) and processed by a clinical expert (3a) to perform RBD diagnosis following a standard manual approach (4). In parallel, actigraphic signals (2b) were processed using machine learning algorithms (3b) to generate an automatic diagnosis of RBD. The study timeline displays the chronological series of recordings performed for each patient, which combined both home and in-clinic evaluations.

#### Clinical classification of patients with vs. without REM sleep behaviour disorder

RBD diagnosis was established based VPSG recordings from two consecutive nocturnal recordings to improve diagnostic power ^28^, and the medical history of each patient. The presence or absence of RBD was established according to standard criteria ^2^. Tonic and phasic muscular activity of REM sleep without atonia (RSWA) were defined according to the international scoring rules ^22^. To have a more refined categorisation, we established a probability score for the presence or absence of RBD in every individual patient, based on both video-PSG recordings of each patient, as follows: a) “definite RBD” (score=1): clear-cut complex dream enactment behaviours and both tonic and phasic RSWA from VPSG; b) “probable RBD” (score=0.75): history of complex dream enactment behaviours and both tonic and phasic RSWA, but not of complex dream enactment behaviours observed at the VPSG; c) “probable no-RBD” (score=0.25): no history of dream enactment behaviour and evidence of only tonic or only phasic RWSA at the VPSG; d) “definite no-RBD” (score=0): no history of dream enactment behaviour and no evidence of RWSA at the VPSG; e) “doubtful RBD” (score=0.5): history of dream enactment behaviour and evidence of only tonic or only phasic RWSA at the VPSG; f) “doubtful no-RBD” (score=0.5): no history of dream enactment behaviour and evidence of both tonic and phasic RWSA at the VPSG.

Each patient was then labelled as “RBD” or “no-RBD” according to the mean of the two individual VPSG scores: “RBD” when the mean score > 0.5, “no-RBD” when the mean score < 0.5. Patients whose score was equal to 0.5 were excluded to ensure that only patients with clear-cut diagnoses were considered. If only one VPSG was available for analyses, the labelling was established based on that one only. Sleep-related respiratory events and periodic limb movements (PLMs) were scored and accounted for (Table 2). We did not find significant different between groups.

We used the STARD checklist when writing our report ^29^.

### Data processing

#### Pre-processing and features extraction

Tri-axial accelerometer signals were segmented for each night, defined as the periods of low illuminance (<200 lux) minus 10 minutes at the beginning and the end. Night-activity tri-axial signals were then combined into a single magnitude vector 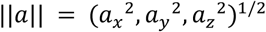, high-pass filtered (4^th^-order Butterworth, cut-off frequency of 0.1Hz), and used to compute features about movement patterns. These features accounted for both (i) the characteristics of isolated, single movement episodes, as well as (ii) global movement patterns over the course of each night (**Fig. 2A,B**).

**Figure 2.**
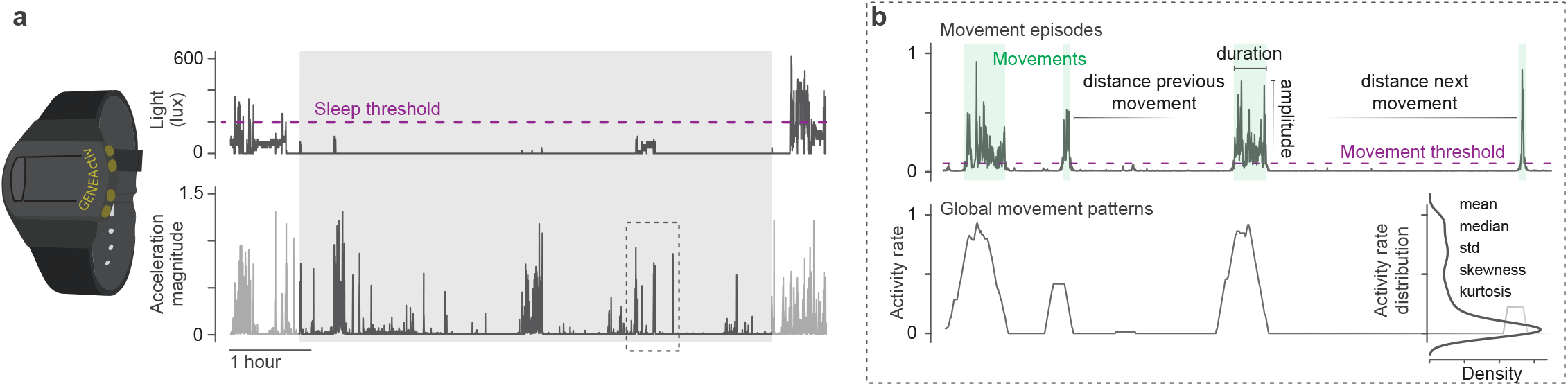
Data processing methodology and feature extraction. **a**, The sleeping period was derived using a light sensor (top), aligned with movement wrist actigraphy recordings (bottom). The night period considered for analyses is shadowed in grey. **b**, Features of nocturnal behaviour were extracted from single movement episodes (top), which characterised behaviour at well-defined isolated times throughout the night, and global movement patterns (bottom).

Movement episodes were identified through thresholding of the acceleration magnitude (threshold = 1*std). We ensured that this value was never below 0.1. Consecutive episodes that were not spaced by at least 1 second were merged into a unique movement event. Each episode was then parameterized by quantifying its duration (short: ≤2s, medium: >2s & ≤10s, long: >10s), magnitude (low: ≤3*movement threshold, high: >3*movement threshold), elapsed time since the previous event, and time to the next (close/clustered: ≤10s, medium: >10s & ≤60s, far/scattered: >60s).

To capture global movement patterns, we additionally computed the rate of activity, defined as the percentage of activity with magnitudes above the predefined threshold within a sliding window (length = 60 seconds, step = 1ms). This activity rate conveys the overall amount of movement throughout the night.

We then computed a series of statistical metrics for each feature such as mean, standard deviation, skewness or kurtosis.

Overall, twenty-nine features were extracted for each night recording (**Supp. Table 1** and **Fig. 2B**). To verify the degree of separability (RBD vs no-RBD patients) captured by the extracted features, we further computed principal component (PC) analysis on this 29-dimensional feature representation.

#### Model construction

We tested several machine learning classification algorithms and compared their performance for discriminating patients with or without RBD.

Prior to model building, a feature selection step was run to reduce the dimensionality of the feature space. Redundant features were first removed if they were not significantly correlated to the subject group (Spearman’s, p>0.05). Least absolute shrinkage and selection operator (LASSO) regularisation was then applied over the retained features: A ranking table was deducted from the subset of features withheld by each LASSO model, computed over 4-fold cross-validation (CV) with 10 repetitions and increasing shrinkage regularisation parameter. Features were ranked based on the percentage of times they were selected by a model. Features selected by less than 10% of the models were discarded.

Classifiers were first built and tested on the data collected during in-lab recordings, from which we identified the best model type and the subset of features to be used for subsequent home recordings. The ability of models to avoid overfitting was determined using a 4-fold CV with class stratification across folds. CV was repeated 100 times to reduce bias in data splitting. We then compared models built from data recorded from either the more affected, less affected, or dominant arm, as well as both arms. In 50% of the patients the dominant arm was the more affected arm.

For home recordings, classifiers were trained on data acquired from three subjects per group (RBD and no-RBD) and tested on all remaining ones (N=20), with 100-time repetition to reduce bias in patient selection. Classification performance was evaluated in terms of accuracy, sensitivity, and specificity. For the home recordings, a receiver operating characteristic (ROC) curve was additionally computed to observe classification performance depending on the class probability threshold.

### Statistical analysis

Differences in population demographics were analysed using the Mann-Whitney U Test, except categorical differences which were investigated using a Chi-squared (χ2) test. The contribution of individual features to help discriminate between RBD and no-RBD patients was evaluated by relating each feature score to the corresponding patient label. Significance was analysed using linear mixed-effects models, with individuals as random effects (to control for repeated measurements per subject). Homoscedasticity was apparent for all models. Comparisons in performance between machine learning models were evaluated using the Mann-Whitney U Test; all results were corrected for multiple comparisons by means of Tukey-Kramer’s correction. All data are reported as mean values ± standard deviation (SD). Stars *,**,*** indicate a significant difference at p < 0.05, p < 0.01 and p < 0.001 respectively.

## RESULTS

### Patients’ population

Twenty-seven PD patients were enrolled in the study. Eighteen patients were labelled as RBD and eight as no-RDB. One patient had to be excluded as their RBD probability score was 0.5, and based on one VPSG recording only (data loss). Patients’ demographic and clinical characteristics are reported in Table 1.

**Table 1.** Patient’s demographic and clinical characteristics.

| Characteristic | RBD<br>(n=18) | no-RBD<br>(n=8) | Difference<br>p-value<sup>1</sup> |
| --- | --- | --- | --- |
| Age (year) | 69.9±8.2 | 63.8±13.9 | 0.29 |
| Sex (M/F) | 15/3 | 4/4 | 0.07<sup>2</sup> |
| Headedness (score) | 74.2±34.9 | 80.6±39.7 | 0.29 |
| More affected side (right/left/symmetrical) | 7/9/2 | 4/2/2 | 0.43 |
| MDS-UPDRS total score | 50.9±23.1 | 46.6±15.4 | 0.59 |
| part I | 8.8±5.1 | 10.2±4.8 | 0.48 |
| part II | 9.7±6.2 | 10.1±4.3 | 0.65 |
| part III (on) | 29.8±14 | 25.6±12.7 | 0.54 |
| part IV | 2.6±2.8 | 0.6±1.2 | 0.07 |
| Hoehn & Yahr stage | 2.0±0.4 | 1.9±0.4 | 0.44 |
| Disease duration (year) | 7.4±5.9 | 4.9±4.9 | 0.26 |
| Presence of motor fluctuations (yes/no) | 1/17 | 1/7 | 0.53<sup>2</sup> |
| Presence of dyskinesias (yes/no) | 10/8 | 2/6 | 0.14<sup>2</sup> |
| Medications |  |  |  |
| levodopa daily equivalent dose (mg) | 589.7±275.6 | 655.3±333.8 | 0.78 |
| benzodiazepines (yes/no) | 5/13 | 1/7 | 0.39<sup>2</sup> |
| melatonin (yes/no) | 0/0 | 0/0 |  |
| antidepressants (yes/no) | 7/11 | 2/6 | 0.49<sup>2</sup> |
| Cumulative illness rating scale - revised (score) | 12.9±3.6 | 21.0±3.6 | 0.62 |
| Cumulative illness rating scale - musculoskeletal (score) | 0.9±0.8 | 1.4±1.1 | 0.25 |
| Parkinson's disease sleep scale (score) | 11.8±7.7 | 14.5±9.8 | 0.43 |
| Pittsburgh sleep quality index (score) | 5.7±2.7 | 6.0±2.1 | 0.75 |
| Epworth sleepiness scale (score) | 6/12 | 4/4 | 0.42<sup>2</sup> |

Clinical scores taken as the average per subject over the entire study. Data are reported as mean ± SD or proportions. <sup>1</sup>Mann-Whitney U Test; <sup>2</sup>Chi-squared test.

**Table 2.** Video-polysomnographic features.

| Characteristic | RBD<br>(n=18) | no-RBD<br>(n=8) | Difference<br>p-value<sup>1</sup> |
| --- | --- | --- | --- |
| Total sleep time (min) | 297.5±69.3 | 316.0±66.0 | 0.56 |
| Sleep onset latency (min) | 12.0±8.2 | 9.5±8.2 | 0.52 |
| REM onset latency (min) | 177.5±61.8 | 201.7±132.0 | 0.74 |
| Wake after sleep onset (min) | 59.5±24.1 | 94.3±66.5 | 0.19 |
| Wake (min) | 70.0±26.9 | 104.0±73.9 | 0.25 |
| Stage N1 (min) | 55.1±27.0 | 70.5±31.6 | 0.29 |
| Stage N2 (min) | 159.2±48.9 | 177.2±67.4 | 0.27 |
| Stage N3 (min) | 60.8±23.0 | 37.0±16.7 | 0.02* |
| REM (min) | 22.5±17.0 | 25.6±14.9 | 0.56 |
| Sleep efficiency (%) | 79.7±7.1 | 75.8±14.5 | 0.8 |
| Apnea index | 35.6±25.3 | 41.8±19.6 | 0.15 |
| Apnea-Hypopnea index | 18.1±14.2 | 16.2±18.9 | 0.7 |
| Respiratory disturbance index | 30.2±26.7 | 33.8±25.9 | 0.81 |
| Periodic limb movement | 10.5±22.4 | 0.9±1.5 | 0.71 |

Video-polysomnographic features taken as the average per subject over the entire study. Data are reported as mean ± SD. <sup>1</sup>Mann-Whitney U Test; \*P < 0.05.

### Clinical validation of RBDAct methodology

#### Extraction of features describing RBD movements and behaviours

We first computed mathematical features that captured nocturnal movement patterns from the acceleration signals. We specifically aimed to account for both (i) the characteristics of single, isolated movement episodes, and (ii) global movement patterns over the course of each night. Overall, twenty-nine features were extracted, for each night (**Fig. 3A**). These were then matched with the corresponding clinical label (RBD vs no-RBD) provided by the clinical expert for training the algorithms.

**Figure 3.**
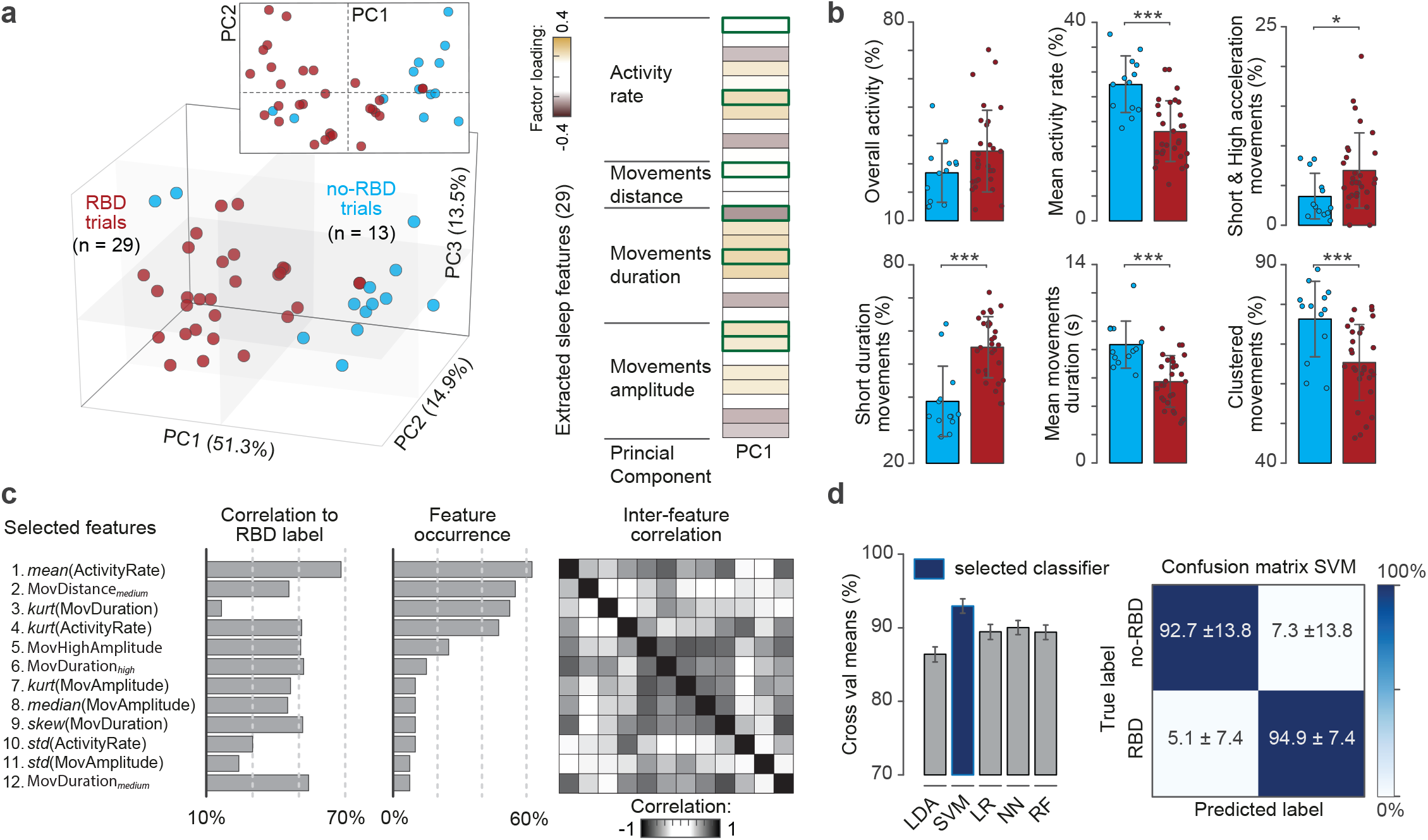
Features capturing RBD movements and behaviours. **a**, Representation of each patient in a low-dimensional feature space (principal components PC1 to PC3). RBD patients indicated in red, and no-RBD patients in cyan. The contribution of each individual feature highlights the movement and behaviours that are most meaningful along PC1. Features outlined in green correspond to those shown in panel b. **b**, Barplots showing group-level differences between RBD and no-RBD patients in gait features identified in a. **c**, A feature selection algorithm identified the most discriminant features between groups using Spearman’s correlation and LASSO regression. **d**, Classification accuracy for the five machine learning algorithms implemented and confusion matrix for the better performing one (SVM). LDA: linear discriminant analysis; SVM: support vector machine; LR: logistic regression; NN: nearest neighbour; RF: random forest.

To verify the capacity of the identified features to capture key differences between RBD and no-RBD patients, we projected the computed 29-dimensional parameterization into a low-dimensional space using PC analysis (**Fig. 3A**). The first 3 PCs explained 79.7% of the overall variance (PC1: 51.3%, PC2: 14.9%; PC3: 13.5%), and highlighted clear differences in space between the two groups. PC1 specifically segregated patients based on the characteristics of movement episodes, based on their duration and magnitude. A closer analysis of the factor loadings of PC1 emphasised that RBD patients exhibited predominantly short, yet high-magnitude movement episodes that were scattered throughout the night, as reflected by the lower mean activity rate and lower percentage of clustered movements (**Fig. 3B**). Additionally, overall nocturnal activity was higher in RBD than no-RBD patients.

We then identified the most meaningful features for classification. A feature selection step was run to extract the ones that maximised the separability between groups. All selected features (N=12) exhibited (i) a high correlation to the patient group (> 10%), (ii) a high occurrence in LASSO regression (> 10%), and (iii) low inter-feature correlation (**Fig. 3C**). As anticipated by the PC analysis, this set of features confirmed that group separability was based on the amount of motor activity throughout the entire night, as well as episode duration and magnitude.

#### In-lab RBDAct classification performance

To automatically discriminate RBD patients using the selected features, we compared the performance of different classification algorithms. All algorithms consistently yielded a high prediction accuracy (mean performance 89.6%), based on the actigraphic recordings acquired during the two nights spent by the patient at the sleep lab. The best performance was achieved by a support vector machine (SVM) model (92.9 ± 8.16% accuracy, 94.9 ± 7.4% sensitivity, 92.7 ± 13.8% specificity; **Fig. 3D**). This model was then retained as the most suitable algorithm to subsequently test home recordings.

We additionally explored if sensor placement had an impact on the features’ ability to capture RBD patterns. We compared the performance of models when the wrist actigraph was worn on the (i) more affected side, (ii) less affected side, (iii) dominant side or (iv) both arms. We only considered patients who exhibited asymmetric motor deficits and wore actigraphic sensors on both arms (N=16 RDB, N=5 no-RBD). Maximum performance was systematically obtained using classifiers that were built on data from the more affected arm, as compared to using the dominant or less affected arm (**Fig. 4**). Placing sensors on both wrists did not improve classification performance.

**Figure 4.**
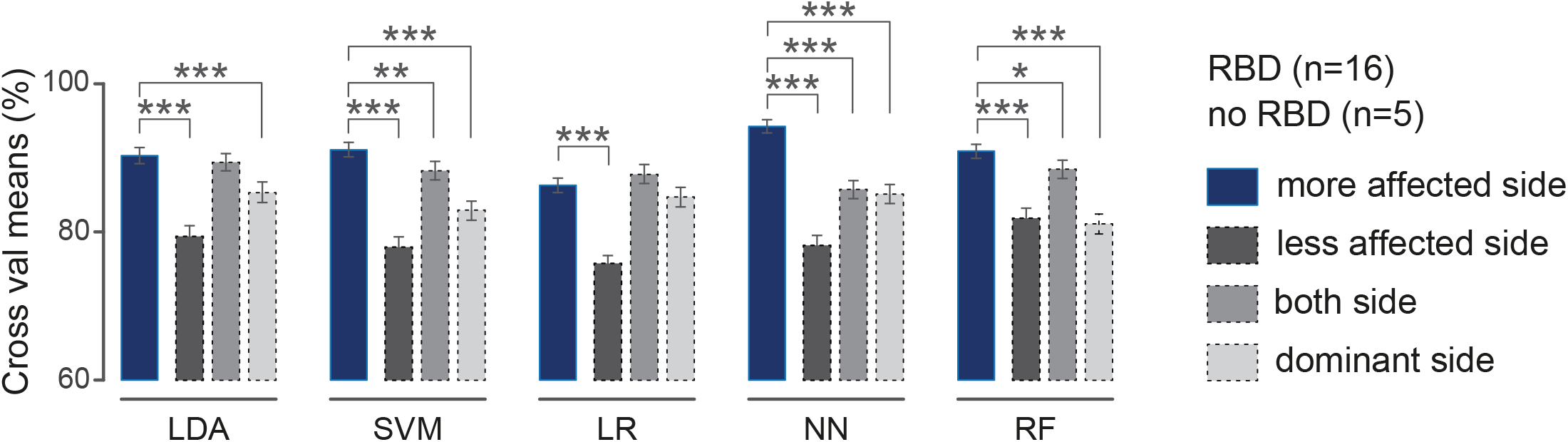
Comparison of classification performance depending on actigraphy sensor position. All algorithms systematically achieved better accuracies when sensors were worn on the most affected side. LDA: linear discriminant analysis; SVM: support vector machine; LR: logistic regression; NN: nearest neighbour; RF: random forest.

### RBDAct performance in home environments

We then tested RBDAct at home. All the patients wore the actigraph during the whole duration of the study (adherence = 100%).

We run our SVM algorithm using the selected features on a 2-week home recording set (**Fig. 5A**). We computed the classification accuracy for each individual night (**Fig. 5B**), and derived a diagnosis from the 2-week probability average to account for daily variability in spontaneous occurrence of RBD movements that would affect classification outcome (**Fig. 5C**). Setting a classification threshold between 0.5 and 0.6 revealed an accuracy of 100% after 7 nights. Progressively increasing the number of nights from 7 to 14, accuracy remained stable between 96 and 100%.

**Figure 5.**
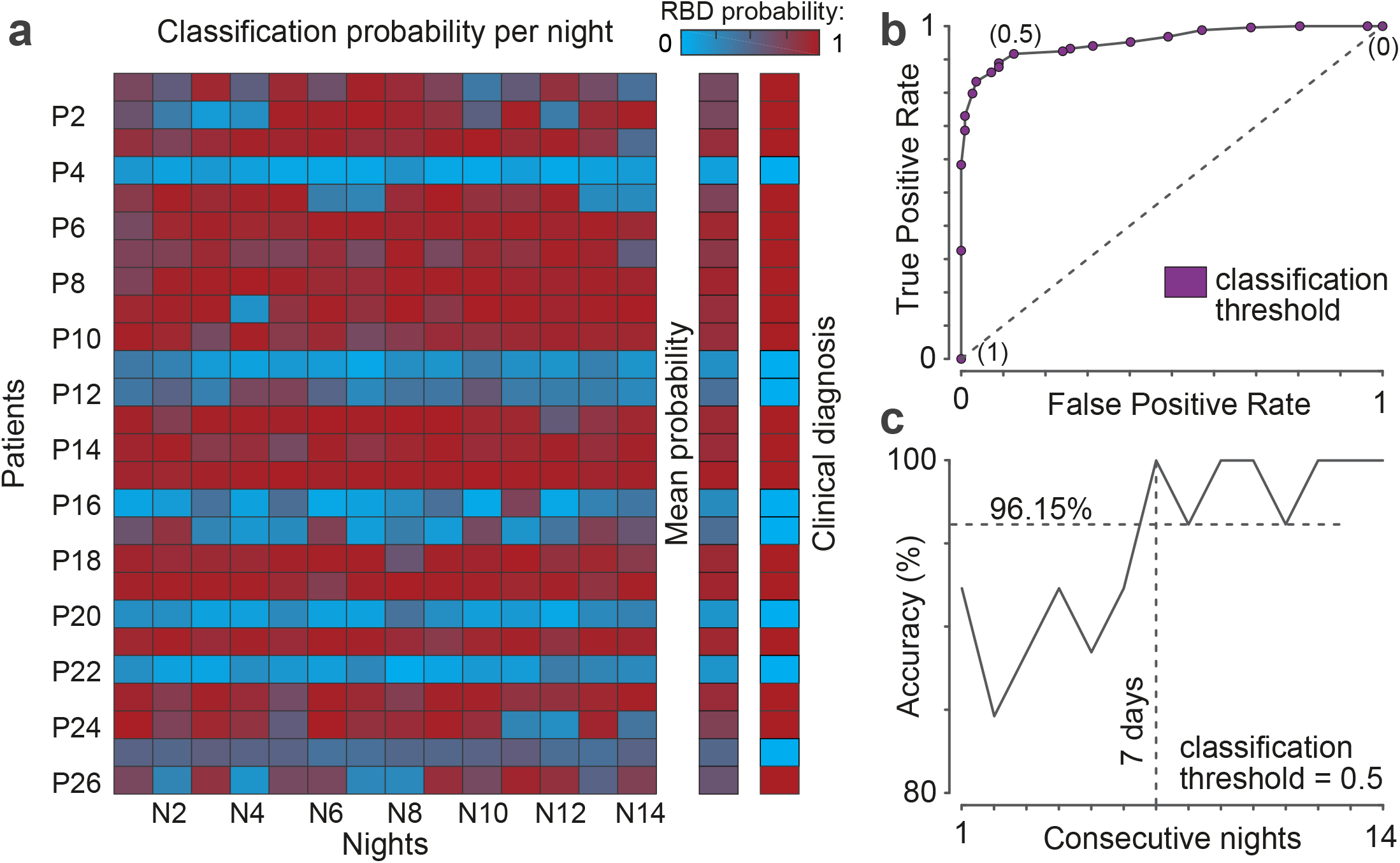
Classification performance during home recordings. **a**, Heatmap of classification probabilities per patient and night (left), and mean probability over the 14-night period aligned to the corresponding clinical diagnosis (right). Probability values range from 0 (cyan, no-RBD) to 1 (red, RBD). **b**, ROC curve to identify the classification threshold (between 0 and 1) that best discriminates RBD vs no-RBD patients across the 14 night period. A threshold of 0.5 was identified as providing the best results. **c**, Changes in classification accuracy when accounting for multiple consecutive nights. Using 7 nights or more lead to performances above 96.15% across patients (threshold = 0.5).

## DISCUSSION

We developed a novel screening tool, termed RBDAct, to automatically identify RBD at home in patients with mild to moderate PD. We first identified features that characterised differences in nocturnal movements and behaviours in RBD vs. no-RBD patients from actigraphic recordings. We then trained various machine learning classification algorithms using in-lab actigraphic data acquired in parallel to VPSG. Classification proved to be highly accurate (92.9 ± 8.16%). Finally, we tested the performance of the best algorithm on a 14-night actigraphic home recording. This out-of-lab validation reached an accuracy of 100% across patients.

### Actigraphic features robustly capture RBD movements and behaviours

RBDAct relied exclusively on accelerometer signals to detect movements and behaviours characteristic of RBD. These have been reported to be qualitatively different from controls during wakefulness, and particularly during arousals and awakenings. RBD movements were reported to be faster, more abrupt, jerky, and violent, both when observed in VPSG or by patients’ bedpartners ^30^. These observations provided the ground for using acceleration as a marker of RBD among the full range of nocturnal movements. Our automated approach confirms these differences from an objective, quantitative standpoint.

Both global night activity patterns, and isolated movement episodes were found to be critical to discriminate between RBD and no-RBD patients, regardless of the analytical methodology employed (i.e. PCA or feature selection algorithms). Features related to global night activity underscored that RBD patients were more active overall, which is in line with VPSG observations ^28 31 32^, and that they exhibited movements that were scattered in time over the course of the night. Instead, patients without RBD moved less frequently and, if they did, their movements were long-lasting and clustered in concise periods of the night. Features related to isolated movement episodes showed that RBD patients exhibit predominantly short, high-magnitude movements compared to no-RBD patients.

From a clinical standpoint, RBD movements and behaviours are expected to cluster intermittently, in correspondence to REM sleep periods. Sleep destructuring in PD ^33 34^, with REM sleep exhibiting a non-nychthemeral distribution, might explain why RBD movements detected by means of actigraphy were found to be spread over the course of the night.

Regardless of cross-patient differences, all tested classification algorithms systematically achieved high performances, confirming the robustness of the identified features to capture key aspects of RBD movements and behaviours. Similar performance was achieved during home recordings, emphasising their stability on multiple observations from the same subject.

### Relevance of the number and location of actigraphic sensors

Maximal classification performance was achieved on average when the sensor was placed on the more affected arm, as compared to the less affected side or the dominant side. This observation suggested that abnormal movements of RBD may be more pronounced on the most affected hemibody. While this may not apply to all individual patients, our experience suggests that the most appropriate *a-priori* placement should be on the most affected arm.

Using two sensors (one per wrist) did not improve the ability to discriminate between RBD vs. no-RBD patients. In some cases, it even worsened prediction accuracy. This suggests that movements of the less affected arm are “less abnormal”, thus reducing the separability between RBD and no-RBD measurements. These observations have compelling practical implications: the ability to restrict recordings to one arm simplifies the setup, increasing comfort and decreasing cost. It certainly accounts for the 100% adherence achieved during home recordings.

### Relevance of the number of nocturnal recordings

Combining measurements from multiple nights proved to be essential to ensure an accurate identification of RBD. In this study, information from VPSG recordings from two nights was necessary to confirm or rule out RBD diagnosis, in a few patients.

An average accuracy of 100% was reached after 7 consecutive nights of actigraphic home recordings. It remained stable between 96-100% when accounting for subsequent nights. Based on these results, we recommend that at least one week of actigraphy data be collected to maximise diagnostic accuracy.

### Limitations and future improvements

Considering the relatively small cohort of patients (18 RBD and 8 non-RBD patients) that were included in the study, the generalisation of our approach for widespread clinical use requires further validations. Our algorithms were trained and tested only on patients with RBD that was secondary to mild or moderate PD. We did not include neither patients with RBD secondary to disorders other than PD, nor patients with isolated RBD.

Similarly, RBDAct did not account for sleep stages in the classification pipeline. Performance may improve by including information about REM or NREM periods. It may be necessary to account for sleep fragmentation and disruption in PD and for the fact that RBD movements may not be exclusively restricted to REM phases, but may also appear at NREM/REM transitions during “covert REM sleep” ^35^ or during “undifferentiated” sleep^34^.

Finally, RBDAct is biased towards identifying patients with RBD characterised by phasic loss of muscle atonia, as only phasic activity can be detected by accelerometers. This is nevertheless more clinically meaningful than tonic RSWA in patient management, to prevent consequences such as injuries to patients or bedpartners. Similarly, RBDAct may have difficulties controlling for RBD-like movements observed on respiratory arousals ^36^ or other sleep-related behaviours. The absence of a control group for these aspects indicates that RBDAct may indeed lead to false positives. Considering that RBDAct is meant to provide a first screening step to guide further in-depth clinical evaluations, our methodology ensures that false negatives are prevented, even at the expense of some false positives.

## CONCLUSION

RBDAct is an innovative technological solution to automatically detect RBD in PD patients. Considering the simplicity of manipulation and affordable price of actigraphy, our approach paves the way for widespread screening of large numbers of patients in ecological environments, both for clinical and research purposes.

Replacing in-lab VPSG with home recordings holds important implications for patients exhibiting severe motor difficulties or dementia, for whom in-lab VPSG can be complex and bothersome. Its potential may also be meaningful for patients who do not have a bed partner. In research, RBDAct would permit large-scale screening and profiling of PD patients during clinical trials. There is a potential for rapid deployment within commercially available technologies, with the advantage of being an automated procedure that is simple to interpret.

RBDAct also has the potential to become a quantitative marker of disease severity, and could be employed to monitor disease progression, to adapt symptomatic treatments, or to evaluate the efficacy of neuro-protective or disease-modifying medications.

Further developments should foresee expanding its applicability in other populations (such as isolated RBD, RBD secondary to other synucleinopathies, or acute, non-degenerative RBD) and for discriminating RBD from other sleep-related behaviours (such as NREM parasomnia, nocturnal epilepsy, arousals from phasic respiratory events).

## Data Availability

Data are available with a granted proposal upon reasonable request.

## Acknowledgments

We would like to thank all the patients who took part in this study. Thanks to Pr. Alain Kaelin-Lang for his strategic support to the Sleep, Awake & Move project, and, together with Dr. Salvatore Galati and Dr. Claudio Staedler, for patient recruitment.

We thank Mr. Paulo-Edson Nunes-Ferreira, Dr. Clara Ferlito, Dr. Sandra Hackethal, Dr. Ninfa Amato, Dr. Serena Caverzasio, Dr. Simona Bonoli, Dr. Lucia Guglielmetti, Dr. Matteo Pereno, Mr. Francesco Mezzanotte for data collection, and Dr. Jihad Louali and Ms. Charlotte Moerman for help with data pre-processing.

We also thank all the colleagues and co-workers who gave scientific, technical or logistic support to the Sleep, Awake & Move project and shared our enthusiasm: Engr. Michele Marazza, Engr. Alessandro Mascheroni, Dr. Francesca Dalia Faraci, Engr. Luigi Fiorillo, Pr. Moustapha Dramé, Mr. Pierluigi Lurà, Mrs. Nicole Vago-Caputo, Mrs. Simona Perrotta, Mr. Giuliano Filippini, Mrs. Yasmin Belloni.

## Contributors

The RBDAct project was an initiative of PLR. PLR, FR, AP and EMM participated in the design of the study. PLR obtained funding to support the study. PLR was responsible of patients’ recruitment and screening and of data collection. FR was the study biostatistician responsible for the statistical analysis. FR, SS, PLR were responsible for processing and analysis of data. FR and SS generated machine learning models. FR was responsible for generating the figures. PLR, AP and EMM were involved in study supervision. All authors were involved in the interpretation of the data and in the manuscript writing. All authors agreed on the content of the manuscript, reviewed drafts, and approved the final version.

## Data availability statement

Data are available with a granted proposal upon reasonable request.

## TABLES

**Supplementary Table 1.** Description of features

| Feature | Description | Category |
| --- | --- | --- |
| Activity | Percentage of overall activity | Global movement patterns |
| Mov number | Total number of movement episodes |  |
| ActivityRate<sub>low</sub> | Percentage of activity rate > 0% and ≤ 30% |  |
| ActivityRate<sub>medium</sub> | Percentage of activity rate > 30% and ≤ 60% |  |
| ActivityRate<sub>high</sub> | Percentage of activity rate > 60% and ≤ 100% |  |
| mean(ActivityRate) | Mean of the activity rate distribution |  |
| median(ActivityRate) | Median of the activity rate distribution |  |
| std(ActivityRate) | Standard deviation of the activity rate distribution |  |
| skew(ActivityRate) | Skewness of the activity rate distribution |  |
| kurt(ActivityRate) | Kurtosis of the activity rate distribution |  |
| MovDistance<sub>close</sub> | Percentage of movement episodes that have distance from the previous/next movement ≤ 10 seconds | Movement episodes |
| MovDistance<sub>medium</sub> | Percentage of movement episodes that have distance from the previous/next movement > 10 and ≤ 60 seconds |  |
| MovDistance<sub>far</sub> | Percentage of movement episodes that have distance from the previous/next movement > 60 seconds |  |
| MovDuration<sub>short</sub> | Percentage of movement episodes that have duration ≤ 2 seconds |  |
| MovDuration<sub>medium</sub> | Percentage of movement episodes that have duration > 2 and ≤ 10 seconds |  |
| MovDuration<sub>long</sub> | Percentage of movement episodes that have duration > 10 seconds |  |
| mean(MovDuration) | Mean of the movement duration distribution |  |
| median(MovDuration) | Median of the movement duration distribution |  |
| std(MovDuration) | Standard deviation of the movement duration distribution |  |
| skew(MovDuration) | Skewness of the movement duration distribution |  |
| kurt(MovDuration) | Kurtosis of the movement duration distribution |  |
| MovHighMagnitude | Percentage of movement episodes with magnitude > 3*movement threshold |  |
| MovHighMagnitudeLowDuration | Percentage of movement episodes that have duration $\leq 2$ seconds and magnitude > 3*movement threshold | |
| MovHighMagnitudeLowDistance | Percentage of movement episodes that have distance from the previous/next movement $\leq 10$ seconds and magnitude > 3*movement threshold | |
| mean(MovMagnitude) | Mean of the movement magnitude distribution |  |
| median(MovMagnitude) | Median of the movement magnitude distribution |  |
| std(MovMagnitude) | Standard deviation of the movement magnitude distribution |  |
| skew(MovMagnitude) | Skewness of the movement magnitude distribution |  |
| kurt(MovMagnitude) | Kurtosis of the movement magnitude distribution |  |

